# Evaluation of the diagnostic value of YiDiXie™-SS, YiDiXie™-HS and YiDiXie™-D in colorectal cancer

**DOI:** 10.1101/2024.08.20.24312286

**Authors:** Xutai Li, Chen Sun, Rui Xiong, Yutong Wu, Huimei Zhou, Zhenjian Ge, Yingqi Li, Wenkang Chen, Wuping Wang, Pengwu Zhang, Shengjie Lin, Siwei Chen, Wei Li, Guoqing Lv, Ling Ji, Yongqing Lai

## Abstract

**Background:** Colorectal cancer poses a severe risk to public health and has a substantial financial impact. Tumor markers such as CEA, CA125, CA19-9, and others, as well as the fecal occult blood test (FOBT), are frequently utilized for colorectal cancer screening and initial diagnosis. However, false-positive results of FOBT, CEA, CA125, and CA19-9 can lead to misdiagnosis and wrong colorectaloscopy, while their false-negative results can lead to missed diagnosis and delayed treatment. Finding practical, affordable, and non-invasive diagnostic techniques is crucial to lowering the false-positive and false-negative rates of FOBT and other indicators. The aim of this study was to evaluate the diagnostic value of YiDiXie™-SS, YiDiXie™-HS and YiDiXie™-D in colorectal cancer.

**Patients and methods:** This study eventually included 916 participants (602 in the malignant group and 314 in the benign group). Serum samples from individuals were obtained and examined using the YiDiXie™ all-cancer detection kit to assess the sensitivity and specificity of YiDiXie™-SS, YiDiXie™-HS and YiDiXie™-D, respectively.

**Results:** The sensitivity of YiDiXie™-SS was 99.0% (95% CI: 97.8% - 99.5%) and its specificity was 63.1% (95% CI: 57.6% - 68.2%). This means that YiDiXie ™ -SS has an extremely high sensitivity and relatively high specificity in colorectal tumors.YiDiXie™-HS has a sensitivity of 93.7% (95% CI: 91.5% - 95.4%) and a specificity of 86.3% (95% CI: 82.1% - 89.7%). This means that YiDiXie™-HS has high sensitivity and specificity in colorectal tumors.YiDiXie™-D has a sensitivity of 82.6% (95% CI: 79.3% - 85.4%) and a specificity of 93.9% (95% CI: 90.7% - 96.1%). This means that YiDiXie ™ -D has relatively high sensitivity and very high specificity in colorectal tumors. YiDiXie ™-SS significantly reduced the false-positive rates of FOBT, CEA, CA125, and CA19-9 with essentially no increase in malignancy leakage. YiDiXie™-HS substantially reduced the false-negative rates of FOBT, CEA, CA125, and CA19-9. YiDiXie™-D significantly reduces the false positive rate of FOBT, CEA, CA125, CA19-9. YiDiXie™-D significantly reduces the false negative rate of FOBT, CEA, CA125, CA19-9 while maintaining a high level of specificity.

**Conclusion:** YiDiXie™-SS has very high sensitivity and relatively high specificity in colorectal tumors.YiDiXie™-HS has high sensitivity and high specificity in colorectal tumors.YiDiXie™-D has relatively high sensitivity and very high specificity in colorectal tumors. YiDiXie ™ -SS significantly reduced false-positive rates for FOBT, CEA, CA125, and CA19-9 with essentially no increase in delayed treatment for colorectal cancer.YiDiXie™-HS substantially reduced false-negative rates for FOBT, CEA, CA125, and CA19-9. YiDiXie™-D can significantly reduce the false-positive rate of FOBT, CEA, CA125 and CA19-9, or significantly reduce the false-negative rate of FOBT, CEA, CA125 and CA19-9 while maintaining a high level of specificity. YiDiXie ™ tests have an important diagnostic value in colorectal cancer, and are expected to solve the problems of “high false-positive rate” and “high false-negative rate” of FOBT, CEA, CA125 and CA19-9.

**Clinical trial number:** ChiCTR2200066840.

## INTRODUCTION

Colorectal cancer is one of the most common malignant tumors. According to the most recent data, there would be 1.92 million new cases of colorectal cancer and 900,000 new deaths worldwide in 2022^1^; the incidence and mortality of colorectal cancer increased by 6.9% and 4.9%, respectively, in 2022 compared to 2020^1-2^. There are stages to colorectal cancer; the disease does not strike suddenly^3^. Furthermore, although early colorectal cancer is asymptomatic, most individuals can be saved from death with early detection.The prognosis for colorectal cancer (CRC) is highly influenced by the cancer’s stage at diagnosis; 5-year survival rates for localized disease are roughly 90%, for regional disease they are 70%, and for distant metastatic CRC they are only 13%^4^. Consequently, colorectal cancer poses a major risk to public health.

Fecal occult blood test (FOBT) and tumor markers such as CEA, CA125, and CA19-9 are widely used in the screening or initial diagnosis of colorectal cancer.On the one hand, FOBT, CEA, CA125, and CA19-9 can produce a large number of false-positive results. In one study, it was shown that the cumulative false-positive rate for FOBT was 16.3%, whereas of all positive FOBTs (2191 out of 4101), false-positive results were obtained in 54.0%^5^. In addition, the false-positive rate of CEA for screening colon cancer was 64%^6^. And also illustrated in another study, 49% of 728 evaluable patients were determined to be false positive for elevated CEA without evidence of recurrent colon cancer^7^. In contrast, the false-positive rate of a single CA19-9 for screening colon cancer is 11%^8^.Patients usually undergo colonoscopy when indicators such as FOBT are positive A false-positive result for an indicator such as FOBT means that the patient undergoes unnecessarily expensive and invasive colonoscopy, and the patient will have to bear the burden of mental anguish The patients will have to bear mental pain, expensive examination costs, examination injuries and other adverse consequences. Therefore, there is an urgent need to find a convenient, economical and non-invasive diagnostic method to reduce the false-positive rate of FOBT and other indicators.

On the other hand, FOBT, CEA, CA125, and CA19-9 can produce a large number of false negative results. A systematic analysis of FOBT as a diagnostic tool noted a false negative rate of 30-70% for FOBT^9-12^. And CEA has a 60% false-negative rate for colon cancer^7^. CA19-9 has a 77% false-negative rate for screening colon cancer^13^. In addition, a single CA125 has a false negative rate of 90% for screening colon cancer^8^. When indicators such as FOBT are negative, patients are usually taken for observation and regular follow up. False-negative results for indicators such as FOBT imply that the malignancy has been underdiagnosed, and will likely lead to delayed treatment, progression of the malignancy, and possibly even development of advanced stages. Patients will thus have to bear the adverse consequences of poor prognosis, high treatment costs, poor quality of life, and short survival. Therefore, there is an urgent need to find a convenient, economical and non-invasive diagnostic method to reduce the false-negative rate of FOBT and other indicators.

Shenzhen KeRuiDa Health Technology Co., Ltd has developed the YiDiXie ™ all-cancer test (henceforth referred to as the “YiDiXie™ test”), an in vitro diagnostic test that can detect multiple cancer types with only 200 microliters of whole blood or 100 microliters of serum each time, based on the discovery of new miRNA tumor markers in serum^14^.YiDiXie ™ test, which can detect multiple cancer types with only 200 microliters of whole blood or 100 microliters of serum per test^14^. The YiDiXie ™ test consists of YiDiXie ™ -HS, YiDiXie ™ -SS, and YiDiXie ™ -D. These three products with very different performance^14^.

The purpose of this study was to evaluate the diagnostic value of YiDiXie™-SS, YiDiXie™-HS and YiDiXie™-D in colorectal cancer.

## PATIENTS AND METHODS

### Study design

This work is a component of the sub-study “Evaluation of the value of the YiDiXie™ test as an adjuvant diagnostic in a variety of tumors” under the SZ-PILOT study (ChiCTR2200066840).

SZ-PILOT is a prospective, observational, single-center study (ChiCTR2200066840). 0.5 ml of the residual serum samples from the subjects who signed a pan-informed permission form for the donation of residual samples at the time of physical examination or admission were collected for use in this investigation.

The study was blinded. The YiDiXie™ test was performed by laboratory workers without knowledge of the individuals’ clinical information. The results were assessed by Coretta laboratory technicians. The clinical professionals evaluating the individuals’ clinical information were ignorant of the YiDiXie™ test results.

The Ethics Committee of Shenzhen Hospital at Peking University approved the study, which was carried out in accordance with the International Conference on Harmonization (ICH) Code of Practice for the Quality Management of Pharmaceutical Clinical Trials and the Declaration of Helsinki.

### Participants

Subjects with colorectal cancer and benign colorectal disease with FOBT, CEA, CA125, and CA19-9 testing data were enrolled separately, and all subjects who met the inclusion criteria were consecutively included.

The study initially included hospitalized patients with “suspected (solid or hematological) malignancy” who completed a pan-informed consent form to donate the remaining samples. Subjects having a postoperative pathological diagnosis of “malignant tumor” were classified as malignant, whereas those with a postoperative pathological diagnosis of “benign disease” were classified as benign. Participants who had unclear pathologic results were excluded from the research. The benign group also included healthy people with colonoscopy results. Some of the samples from the malignant group and samples from healthy examinees in the benign group were used in other previous studies by our group^14^.

Subjects who failed the serum sample quality test prior to the YiDiXie™ test were excluded from this study. Refer to the previous article of this subject group for specific enrollment and exclusion ^14^.

### Sample collection, processing

The serum samples utilized in this investigation were obtained from serum leftover from a routine consultation, eliminating the need for extra blood sampling. The Medical Laboratory took about 0.5 cc of serum from participants and kept it at -80°C for the YiDiXie™ test.

### The YiDiXie test

Shenzhen KeRuiDa Health Technology Co., Ltd created and manufactures the YiDiXie™ all-cancer detection kit, an in vitro diagnostic kit for fluorescent quantitative PCR apparatus, which is used to perform the YiDiXie ™ test. To ascertain whether cancer is present in the subject’s body, it measures the expression levels of many miRNA biomarkers in the serum.^14^ It maintains the specificity and increases the sensitivity of a wide range of malignancies by integrating these independent assays in a contemporaneous testing format and predefining suitable criteria for each miRNA biomarker to guarantee that each miRNA marker is highly specific.^14^

YiDiXie ™ -HS, YiDiXie ™ -SS, and YiDiXie ™ -D are the three distinct tests that make up the YiDiXie ™ test.^14^ Of these, YiDiXie™-HS was created with attention to both specificity and sensitivity.^14^ YiDiXie ™ -SS greatly enhanced the quantity of miRNA assays to get very high sensitivity for every clinical stage of every kind of malignant tumor.^14^ With YiDiXie™-D, the diagnostic threshold of individual miRNA testing is significantly raised, resulting in very good specificity for all forms of cancer.^14^

Perform the YiDiXie™ test according to the instructions of the YiDiXie™ Whole Cancer Test. Refer to our previous work for detailed procedure^14^.

The laboratory technicians of Shenzhen KeRuiDa Health Technology Co., Ltd examined the raw test results and determined that the YiDiXie™ test had either “positive” or “negative” results^14^.

### Extraction of clinical data

The clinical, pathological, laboratory, and imaging data for this investigation were obtained from the individuals’ inpatient medical records or physical examination reports. Clinical staging was performed by trained physicians using the AJCC staging manual (7th or 8th edition)^15-16^.

### Statistical analyses

Demographic and baseline variables were recorded using descriptive statistics. For categorical variables, the number and percentage of subjects in each category were calculated; for continuous variables, the total number of subjects (n), mean, standard deviation (SD) or standard error (SE), median, first quartile (Q1), third quartile (Q3), minimum and maximum values were calculated. The Wilson (score) technique was used to compute the 95% confidence intervals (CIs) for several indicators.

## RESULTS

### Participant disposition

This study involved a total of 916 participants (602 malignant cases and 314 benign cases). The demographic and clinical characteristics of the 916 study participants are shown in Table 1.

**Table 1.** Participants’ demographic and clinical manifestation.

|  | Malignant (n =602) | Benign (n =314) | Total (N = 916) |
| --- | --- | --- | --- |
| Age, years |  |  |  |
| Mean (SD) | 63.1 (10.95) | 47.1 (17.66) | 56.3 (16.25) |
| Median (Q1,Q3) | 65 (56, 70) | 53 (27, 60) | 58 (52, 66) |
| Min, max | 25, 81 | 21, 76 | 21, 81 |
| Age, group, n (%) |  |  |  |
| < 50 | 2 (5.4) | 12 (48.0) | 14 (22.6) |
| ≥ 50 | 35 (94.6) | 13 (52.0) | 48 (77.4) |
| < 65 | 18 (48.6) | 22 (88.0) | 40 (64.5) |
| ≥ 65 | 19 (51.4) | 3 (12.0) | 22 (35.5) |
| Sex, n (%) |  |  |  |
| Female | 17 (45.9) | 19 (76.0) | 357 (39.0) |
| Male | 20 (54.1) | 6 (24.0) | 559 (61.0) |
| Body mass index (kg/m2) |  |  |  |
| n | 37 | 23 | 60 |
| Mean (SD) | 22.1 (2.32) | 22.1 (3.47) | 22.1 (2.79) |
| Median (Q1,Q3) | 21.5 (20.4, 22.9) | 22.2 (20.6, 24.2) | 21.9 (20.2, 24.0) |
| Min, max | 18.6 28.2 | 15.6 28.5 | 15.6 28.5 |
| Body mass index category, n (%) |  |  |  |
| Underweight | 0 (0) | 4 (16.0) | 4 (6.5) |
| Benign | 28 (75.7) | 12 (48.0) | 40 (64.5) |
| Overweight | 8 (21.6) | 5 (20.0) | 13 (21.0) |
| Obese | 1 (2.7) | 2 (8.0) | 3 (4.8) |
| Missing | 0 (0) | 2 (8.0) | 2 (3.2) |
| AJCC clinical stage |  |  |  |
| Stage I | 11 (29.7) |  | 11 (29.7) |
| Stage II | 9 (24.3) |  | 9 (24.3) |
| Stage III | 6 (16.2) |  | 6 (16.2) |
| Stage IV | 10 (27.0) |  | 10 (27.0) |
| Missing | 1 (2.7) |  | 1 (2.7) |

Q1,Q3, first quartile, third quartile; SD, standard deviation.

Demographic and clinical variables were similar between the two research groups (Table 1). The average (standard deviation) age was 56.3 (16.25) years, with 39.0% (357/916) being female.

### Diagnostic performance of YiDiXie™-SS

As shown in Table 2, the sensitivity of YiDiXie™ -SS was 99.0% (95% CI: 97.8% - 99.5%) and its specificity was 63.1% (95% CI: 57.6% - 68.2%). This means that YiDiXie ™ -SS has very high sensitivity and relatively high specificity in colorectal tumors.

**Table 2.** Performance of YiDiXie™-SS.

| Table 2. Performance of YiDiXie™-SS |  |  |  |
| --- | --- | --- | --- |
|  | Malignant | Benign | Total |
|  | 602 | 314 | 916 |
| Positive | 596 | 116 | 712 |
| Negative | 6 | 198 | 204 |
| SEN = $596/602 = 99.0\%$ (97.8% - 99.5%) SPE = $198/314 = 63.1\%$ (57.6% - 68.2%) | | | |
| SEN, Sensitivity. SPE, Specificity. Two-sided 95% Wilson confidence intervals were calculated. |  |  |  |

### Diagnostic performance of YiDiXie™-HS

As shown in Table 3, the sensitivity of YiDiXie™ -HS was 93.7% (95% CI: 91.5% - 95.4%) and its specificity was 86.3% (95% CI: 82.1% - 89.7%). This means that YiDiXie ™ -HS has high sensitivity and specificity in colorectal tumors.

**Table 3.** Performance of YiDiXie™-HS.

| Table 3. Performance of YiDiXie™-HS |  |  |  |
| --- | --- | --- | --- |
|  | Malignant | Benign | Total |
|  | 602 | 314 | 916 |
| Positive | 564 | 43 | 607 |
| Negative | 38 | 271 | 309 |
| SEN = $564/602 = 93.7\%$ (91.5% - 95.4%) SPE = $271/314 = 86.3\%$ (82.1% - 89.7%) | | | |
| SEN, Sensitivity. SPE, Specificity. Two-sided 95% Wilson confidence intervals were calculated. |  |  |  |

### Diagnostic performance of YiDiXie™-HS

As shown in Table 4, the sensitivity of YiDiXie™ -D was 82.6% (95% CI: 79.3% - 85.4%) and its specificity was 93.9% (95% CI: 90.7% - 96.1%). This means that YiDiXie ™ -D has relatively high sensitivity and very high specificity in colorectal tumors.

**Table 4.** Performance ofYiDiXie™-D.

|  | Malignant | Benign | Total |
| --- | --- | --- | --- |
|  | 602 | 314 | 916 |
| Positive | 497 | 19 | 516 |
| Negative | 105 | 295 | 400 |

SEN = $497/602 = 82.6\%$ (79.3% - 85.4%)
SPE = $295/314 = 93.9\%$ (90.7% - 96.1%)
SEN, Sensitivity. SPE, Specificity. Two-sided 95% Wilson confidence intervals were calculated.

### Diagnostic Performance of YiDiXie™-SS in FOBT, CEA, CA125, CA19-9-positive or negative patients

In order to solve the difficult problem of high false-positive rate of FOBT, CEA, CA125 and CA19-9, YiDiXie ™ -SS was applied to FOBT, CEA, CA125 and CA19-9 positive patients.

As shown in Table 5, YiDiXie™-SS significantly reduced the false-positive rates of FOBT, CEA, CA125, and CA19-9 with essentially no increase in malignancy leakage.

**Table 5.** Performance of different Items.

| Items | Total N | Positive | Sensitivity % (95% CI)<sup>a</sup> | Total N | Negative | Specificity % (95% CI)<sup>b</sup> |
| --- | --- | --- | --- | --- | --- | --- |
| YiDiXie™-SS in FOBT-positive | 210 | 209 | 99.5% (97.4% - 100%) | 22 | 12 | 54.5% (34.7% - 73.1%) |
| YiDiXie™-HS in FOBT-negative | 91 | 80 | 87.9% (79.6% - 93.1%) | 24 | 21 | 87.5% (69.0% - 95.7%) |
| YiDiXie™-D in FOBT-positive | 210 | 156 | 74.3% (68.0% - 79.7%) | 22 | 20 | 90.9% (72.2% - 98.4%) |
| YiDiXie™-D in FOBT-negative | 91 | 69 | 75.8% (66.1% - 83.5%) | 24 | 21 | 87.5% (69.0% - 95.7%) |
| YiDiXie™-SS in CEA-positive | 200 | 199 | 99.5% (97.2% - 100%) | 20 | 11 | 55.0% (34.2% - 74.2%) |
| YiDiXie™-HS in CEA-negative | 367 | 340 | 92.6% (89.5% - 94.9%) | 293 | 254 | 86.7% (82.3% - 90.1%) |
| YiDiXie™-D in CEA-positive | 200 | 174 | 87.0% (81.6% - 91.0%) | 20 | 17 | 85.0% (64.0% - 94.8%) |
| YiDiXie™-D in CEA-negative | 367 | 294 | 80.1% (75.7% - 83.9%) | 293 | 277 | 94.5% (91.3% - 96.6%) |
| YiDiXie™-SS in CA125-positive | 34 | 34 | 100% (89.8% - 100%) | 5 | 2 | 40.0% (7.1% - 76.9%) |
| YiDiXie™-HS in CA125-negative | 513 | 480 | 93.6% (91.1% - 95.4%) | 192 | 163 | 84.9% (79.1% - 89.3%) |
| YiDiXie™-D in CA125-positive | 34 | 30 | 88.2% (73.4% - 95.3%) | 5 | 4 | 80.0% (37.6% - 99.0%) |
| YiDiXie™-D in CA125-negative | 513 | 420 | 81.9% (78.3% - 85.0%) | 192 | 174 | 90.6% (85.7% - 94.0%) |
| YiDiXie™-SS in CA19-9-positive | 72 | 72 | 100% (94.9% - 100%) | 9 | 6 | 66.7% (35.4% - 87.9%) |
| YiDiXie™-HS in CA19-9-negative | 494 | 459 | 92.9% (90.3% - 94.9%) | 286 | 245 | 85.7% (81.1% - 89.3%) |
| YiDiXie™-D in CA19-9-positive | 72 | 70 | 97.2% (90.4% - 99.5%) | 9 | 9 | 100% (70.1% - 100%) |
| YiDiXie™-D in CA19-9-negative | 494 | 397 | 80.4% (76.6% - 83.6%) | 286 | 267 | 93.4% (89.9% - 95.7%) |

CI, confidence interval. <sup>a,b</sup> Two-sided 95% Wilson confidence intervals were calculated.

### Diagnostic Performance of YiDiXie™-HS in FOBT, CEA, CA125, CA19-9-negative patients

In order to solve the difficult problem of high false-negative rate of FOBT, CEA, CA125, and CA19-9, YiDiXie™-HS was applied to FOBT, CEA, CA125, and CA19-9 negative patients.

As shown in Table 5, YiDiXie™-HS substantially reduced the false-negative rates of FOBT, CEA, CA125, and CA19-9.

### Diagnostic Performance of YiDiXie™-D in FOBT, CEA, CA125, CA19-9-positive or negative patients

In order to further reduce the false-positive rate of FOBT, CEA, CA125, and CA19-9, therefore, YiDiXie™-D, which has relatively high sensitivity and very high specificity, was applied to FOBT, CEA, CA125, and CA19-9 positive patients.

As shown in Table 5, YiDiXie™-D substantially reduced the false-positive rates of FOBT, CEA, CA125, and CA19-9 while maintaining high specificity.

### Diagnostic Performance of YiDiXie™-D in FOBT, CEA, CA125, CA19-9-negative patients

In order to reduce the false-negative rate of FOBT, CEA, CA125, and CA19-9 while maintaining a high degree of specificity, YiDiXie™-D, which has a relatively high sensitivity and very high degree of specificity, was applied to FOBT, CEA, CA125, and CA19-9 negative patients.

As shown in Table 5, YiDiXie™-D significantly reduced the false-negative rates of FOBT, CEA, CA125, and CA19-9 while maintaining high specificity.

As shown in Table 5, YiDiXie™-D substantially reduced the false-positive rates of FOBT, CEA, CA125, and CA19-9.

## DISCUSSION

### Clinical significance of YiDiXie™-SS in patients with positive FOBT and other indicators

The YiDiXie™ test consists of three tests with very different characteristics: YiDiXie™-HS, YiDiXie™-SS and YiDiXie™-D).^14^ YiDiXie™-HS combines high sensitivity and high specificity^14^, while YiDiXie™-SS has very high sensitivity for all malignant tumor types, but slightly lower specificity^14^. YiDiXie™-D has very high specificity for all malignant tumor types, but lower sensitivity^14^.

In patients with positive FOBT and other signs, additional diagnostic tests are required for both sensitivity and specificity. evaluating the contradiction between sensitivity and specificity entails evaluating the “danger of malignant tumors being missed” against the “danger of benign tumors being misdiagnosed”. In general, when FOBT and other signs are positive, colonoscopy is used instead of drastic surgery. As a result, false positives like FOBT do not have catastrophic repercussions such as substantial surgical trauma, organ removal, or loss of function. Thus, among patients with positive FOBTs, the “risk of malignant tumor underdiagnosis” is significantly greater than the “risk of benign tumor misdiagnosis”.Therefore, YiDiXie™-SS, which has a very high sensitivity but a slightly lower specificity, was chosen to reduce the false positive rate of FOBT and other indicators.

As shown in Table 5, YiDiXie™-SS significantly reduced the false-positive rates of FOBT, CEA, CA125, and CA19-9 with essentially no increase in malignancy leakage.

According to the aforementioned findings, YiDiXie™-SS virtually eliminates the possibility of misinterpreting colonoscopy results for benign colorectal illness while virtually halving the number of malignant tumors overlooked. Stated differently, YiDiXie™-SS significantly lessens the psychological distress, costly testing expenses, testing-related injuries, and other unfavorable outcomes for patients with false-positive FOBT, CEA, CA125, and CA19-9 without actually increasing the number of postponed treatments for malignant tumors. As a result, YiDiXie™-SS offers a wide range of application potential, significant clinical value, and effectively satisfies clinical needs.

### Clinical significance of YiDiXie™-HS in patients with negative FOBT and other indicators

The sensitivity and specificity of additional diagnostic techniques are critical for patients with negative FOBT and other signs. Weighing the conflict between the “harm of malignant tumor underdiagnosis” and the “harm of benign disease misdiagnosis” is the equivalent of weighing the conflict between sensitivity and specificity. Increased false-negative rates translate into an increased number of malignant tumors being underdiagnosed, which delays treatment, causes the tumor to develop, and can even reach advanced stages. Patients will consequently have to deal with the negative effects of a bad prognosis, a brief surviving time, a low quality of life, and expensive therapy. A higher false-positive rate translates into more benign diseases being misdiagnosed, which will needlessly result in an invasive and costly colonoscopy. Patients must therefore deal with the effects of psychological distress, costly testing, and injury. Therefore, YiDiXie™-HS, with its high sensitivity and specificity, was chosen to reduce the false negative rate of FOBT and other indicators.

As shown in Table 5, YiDiXie™-HS substantially reduced the false-negative rates of FOBT, CEA, CA125, and CA19-9.

YiDiXie™-HS significantly minimizes false-negative diagnoses of malignant tumors using FOBT and other markers, as shown in the above results. YiDiXie™-HS significantly minimizes the poor prognosis, high treatment costs, poor quality of life, short survival duration, and other unfavorable repercussions of patients with false-negative missed diagnosis of malignant tumors by FOBT or other indicators. YiDiXie™-HS addresses clinical needs and has a wide range of applications.

### Clinical significance of YiDiXie™-D in patients with colorectal tumor

For patients with colorectal tumors, YiDiXie™-D, which has relatively high sensitivity and very high specificity, can be used to further reduce the false-positive rates of FOBT, CEA, CA125, and CA19-9 or to significantly reduce their false-negative rates while maintaining high specificity.

As shown in Table 5, YiDiXie™-D substantially reduced the false-positive rates of FOBT, CEA, CA125, and CA19-9. YiDiXie ™ -D significantly reduced the false-negative rates of FOBT, CEA, CA125, and CA19-9 while maintaining a high degree of specificity.

The above results imply that YiDiXie™-D further reduces the risk of incorrectly performed colorectoscopies. Therefore, YiDiXie™-D meets the clinical needs well and has important clinical significance and wide application prospects.

### The YiDiXie™ test promises to address 2 challenges in colorectal cancer

First of all, YiDiXie™-SS relieves non-essential job pressure for gastrointestinal endoscopists, allowing for rapid identification and treatment of malignant tumors that might otherwise be delayed. When signs like FOBT are positive, the patient usually gets a colonoscopy. The quantity of gastrointestinal endoscopists directly influences the timely completion of these colonoscopies. In many parts of the world, appointments are planned months, if not years in advance. This obviously delays the treatment of malignancy cases among them, therefore it is not uncommon for patients with positive FOBT and other symptoms who are waiting for a colonoscopy to have malignancy progression or even distant metastases. As shown in Table 5, YiDiXie™-SS significantly reduced the false-positive rates of FOBT, CEA, CA125, and CA19-9 with essentially no increase in malignancy leakage. As a result, YiDiXie™-SS can significantly relieve gastrointestinal endoscopists of non-essential workloads and facilitate the timely diagnosis and treatment of colorectal cancer or other diseases that would otherwise be delayed.

Second, YiDiXie ™ -HS can greatly reduce the risk of colorectal cancer underdiagnosis. When indicators such as FOBT are negative, colorectal cancer is usually ruled out for the time being. The high rate of false-negative FOBT and other indicators leads to delayed treatment for a large number of colorectal cancer patients. As shown in Table 5, YiDiXie ™ -HS substantially reduced the false-negative rates of FOBT, CEA, CA125, and CA19-9. Therefore, YiDiXie ™ -HS significantly reduces the probability of false-negative diagnosis of malignant tumors by FOBT and other indicators, and facilitates timely diagnosis and treatment of colorectal cancer patients who would otherwise be delayed in treatment.

Again, YiDiXie ™ -D is expected to further address the challenges of “high false positive rate” and “high false negative rate”. As shown in Table 5, YiDiXie™-D substantially reduces the false-positive rates of FOBT, CEA, CA125, and CA19-9, or significantly reduces their false-negative rates while maintaining high specificity. Thus, YiDiXie ™ -D further reduces the risk of incorrectly performed colorectoscopies.

Final, The YiDiXie™ test provides “just-in-time diagnosis” for colorectal cancer patients. The YiDiXie™ test is non-invasive and requires only a small sample of blood, allowing patients to complete the diagnostic process from home. A YiDiXie™ test requires only 20 microliters of serum, which is equivalent to approximately one drop of whole blood. One drop of whole blood is approximately 50 microliters, which provides 20-25 microliters of serum^14^.Taking into account the pre-test sample quality assessment test and 2-3 repetitions, 0.2 ml of whole blood is sufficient for the YiDiXie™ test^14^. The 0.2 ml of finger blood can be collected at home by the average patient using a finger blood collection needle, eliminating the need for venous blood collection by medical personnel and allowing patients to complete the diagnostic process non-invasively without leaving their homes^14^.

On the other hand, the diagnostic capacity of the YiDiXie ™ test is virtually unlimited. Figure 1 shows the basic flowchart of the YiDiXie ™ test, which shows that the YiDiXie™ test not only does not require a doctor or medical equipment, but also does not require medical personnel to collect blood. As a result, the YiDiXie™ test is completely independent of the number of medical personnel and facilities, and has a virtually unlimited test capacity. In this way, the YiDiXie ™ test enables “just-in-time diagnosis” of colorectal cancer patients without the need for patients to wait anxiously for an appointment.

**Figure 1.**
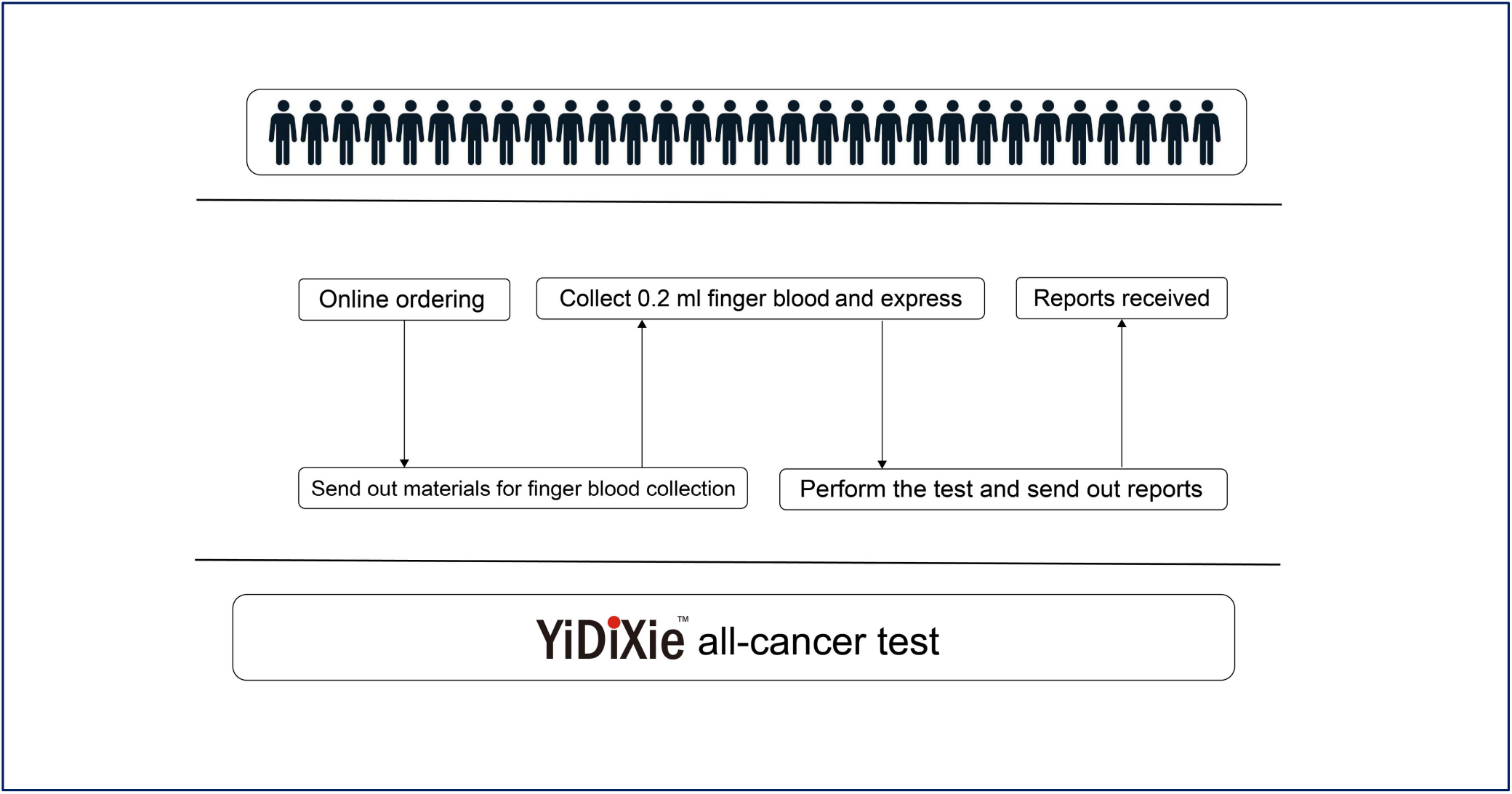
Basic flowchart of the YiDiXie™ test.

In short, The YiDiXie™ test is a valuable diagnostic tool for colorectal cancer, addressing the issues of “high false-positive rate of FOBT and other indicators” and “high false-negative rate of FOBT and other indicators”.

### Limitations of the study

First, this study had a small number of cases and future clinical studies with larger sample sizes are needed for further evaluation.

Second, the present study was an inpatient malignant tumor case-benign tumor control study, and future cohort studies in the natural population of brain tumors are needed for further evaluation.

Final, the current study was a single-center study, which may have led to some degree of bias in the results of this study. Multicenter studies are needed to further evaluate this in the future.

## CONCLUSION

YiDiXie ™ -SS has very high sensitivity and relatively high specificity in colorectal tumors.YiDiXie ™-HS has high sensitivity and high specificity in colorectal tumors.YiDiXie ™ -D has relatively high sensitivity and very high specificity in colorectal tumors. YiDiXie ™ -SS significantly reduced false-positive rates for FOBT, CEA, CA125, and CA19-9 with essentially no increase in delayed treatment for colorectal cancer.YiDiXie ™ -HS substantially reduced false-negative rates for FOBT, CEA, CA125, and CA19-9. YiDiXie ™ -D can significantly reduce the false-positive rate of FOBT, CEA, CA125 and CA19-9, or significantly reduce the false-negative rate of FOBT, CEA, CA125 and CA19-9 while maintaining a high level of specificity. YiDiXie™ tests have an important diagnostic value in colorectal cancer, and are expected to solve the problems of “high false-positive rate” and “ high false-negative rate “ of FOBT, CEA, CA125 and CA19-9.

## Data Availability

All data produced in the present study are contained in the manuscript.

